# Predictive Model with Analysis of the Initial Spread of COVID-19 in India

**DOI:** 10.1101/2020.05.02.20088997

**Authors:** Shinjini Ghosh

**Affiliations:** Massachusetts Institute of Technology 77 Massachusetts Avenue, MA 02139, USA

**Author notes:** Corresponding author; www.mit.edu/~shinghos/.

**Keywords:** COVID-19, India, Modeling, Predictive System, Statistical Analysis

## Abstract

The Coronavirus Disease 2019 (COVID-19) has currently ravaged through the world, resulting in over *three million* confirmed cases and over *two hundred thousand* deaths, a complete change in daily life as we know it, worldwide lock-downs, travel restrictions, as well as heightened hygiene measures and physical distancing. Being able to analyse and predict the spread of this epidemic-causing disease is hence of utmost importance now, especially as it would help in the reasoning behind important decisions drastically affecting countries and their people, as well as in ensuring efficient resource and utility management. However, the needs of the people and specific conditions of the spread are varying widely from country to country. In this article, we have conducted an in-depth analysis of the spread of COVID-19 in India, patient statistics, as well as proposed a mathematical prediction system that has succeeded in predicting the following day’s number of cases with 83% accuracy, ever since the first COVID-19 case was declared in India.

## 1. Introduction

*COVID-19*. Coronavirus Disease 2019 (COVID-19) which originated from the “SARS-CoV-2” virus has led to an ongoing pandemic which has over 2,959,929 confirmed cases and 202,733 confirmed deaths over 213 countries, areas, and territories (WHO)[1] as of April 27, 2020.

*COVID-19 in India*. As of April 28, 2020, India reported a total of 29, 974 COVID-19 cases, including 111 foreign nationals, across 32 states and union territories (MoHFW)[2]. On January 30, 2020, the first laboratory confirmed case of COVID-19 in India was reported in Kerala, a southwestern state (WHO)[3]. Since then, cases have been on the rise all over India and several measures have been implemented to help curb the spread, including suspension of commercial passenger aircraft since March 22, 2020, and imposing a 21-day countrywide lockdown on March 25, 2020, which later got extended until May 3, 2020. On May 01, 2020, the lockdown got further extended by another fortnight. As of March 28, India had 909 confirmed cases of COVID-19, just a month before April 28’s figure of 29, 974, which is almost 33 times higher.

The present article delves into more specifics of the spread of COVID-19 in India, conducting a statistical analysis of the affected patients, as well as proposing a new mathematical model to predict the spread of the disease in the country. Section 2 outlines how the data has been collected, Section 3 discusses the patient-level statistical analysis and compares that with world trends, and Section 4 looks at the details of the modeling and prediction system. The results have then been discussed and analysed thoroughly in Section 5. Concluding remarks and areas for future work are in Section 6.

## 2. Data Collection

The data analysed in this paper includes the official counts as released by MoHFW, hundreds of news reports, the COVID-19 India API, as well as volunteer-collected de-identified open source data until the date of April 26, 2020.

## 3. Patient-level Analysis

In this section, we analyse various demographic and other factors of the COVID-19 affected patients in India, and contrast and compare that with world trends.

Age counts of 2339 patients were received, and the mean age was found to be 38.54 years, with a standard deviation of 17.22 years.

Out of the 5313 affected people whose sex was identified, there were 3547 males (66.76%) and 1766 females (33.24%). This is somewhat skewed when compared globally to around 30 other countries, most of whom report males to have only slightly > 50% cases on average (Global Health 5050, WHO).

As of April 26, 2020, there were 26, 917 cases officially confirmed in India, with 5914 (21.97%) of them having recovered, 20177 (74.96%) active cases and 826 (3.07%) deceased. Of the cases with an outcome, 87.74% have recovered, somewhat higher than the current global average of 81% (Worldometers)[4].

Of the affected patients whose status was tracked, an outcome was obtained in a mean time of 10.69 days, with great variance observed — the mean recovery time (time between hospitalization and official status of recovery) was found to be 14.47 days and the mean death time (time between hospitalization and death) was found to be a mere 3.18 days. The recovery rate is at par with CDC guidelines and WHO statements that mention it may take the body up to 2 weeks to recover from the illness, and up to 6 weeks for severe or critical cases. However, the mean time for death is drastically lower than the WHO mentioned figure of 2 – 8 weeks.

We obtained notes regarding transmission for 3273 patients (mostly those infected towards the initial days of spread in India, after which tracking became much more complicated), out of whom 712 patients had travel history, 194 patients were family member transmissions, and an additional 578 patients were direct contacts of another infected person. Furthermore, 911 patients had attended mass gatherings with other infected people.

Out of the 2339 patients whose age data we received, the distribution was observed as in Table 1. We see a sharp contrast in this distribution when compared with other countries / regions of the world. In Italy, about 71% of individuals affected with COVID-19 was over 50 years old (Statista)[5] but in India, that figure is only 27.53%. Only 8.1% of COVID-19 affected patients in China were in their 20’s, versus around 23% of Indians. We also notice that for the age distribution of adults among the COVID-19 confirmed cases in India, the number of people in each age bucket is steadily decreasing, with the largest number of adults infected being the youngest ones — a departure from the world trend.

**Table 1:** Age Distribution among Infected Patients

| Age Range | Number of Patients | Fraction of Total |
| --- | --- | --- |
| $\leq 18$ | 217 | 9.28% |
| 18 – 29 | 558 | 23.86% |
| 30 – 39 | 534 | 22.83% |
| 40 – 49 | 386 | 16.50% |
| 50 – 59 | 311 | 13.30% |
| 60 – 69 | 237 | 10.13% |
| 70 – 79 | 73 | 3.12% |
| 80 – 89 | 18 | 0.77% |
| $\geq 90$ | 5 | 0.21% |

## 4. Modeling and Predicting the Number of Daily Cases

### 4.1 Introduction

A very important aspect of attempting to control this ongoing pandemic and save lives is to model the spread that has occurred so far, and to be able to use that to predict future spread and number of cases. There has been extensive ongoing research in this aspect, catering to the needs of various countries. In the rest of this article, we propose a model and prediction system that is developed very specifically with India in mind, and currently offers excellent predictions.

#### Background Research

There are multiple models attempting to predict the spread of coronavirus in the world currently. These notably include the UPenn CHIME model [6], a discrete-time SIR (Susceptible-Infected-Recovered) modeling of infections/recovery, which as shown in the following paragraph, fails on predicting the spread of COVID-19 in India. Another SIR based model being used widely includes Health Catalyst [7]. Non-SIR models performing statistical/mathematical modeling are also evolving, such as a Gauss error function and Monte Carlo simulation [8] for Italy and other statistical models [9] in the USA — nonetheless, none of them satisfactorily predicting the spread yet, especially when applied to India.

#### Background Research for India

There have been limited research in modeling and predicting the spread of COVID-19 in India so far, owing both to the ongoing nature of the pandemic and limited availability of data. In an article published in early April 2020 [10], we see the usage of the currently famous SIR Model, and a prediction that India will reach a final epidemic size of around 13, 000 by the end of May 2020. Needless to say, this is a drastic difference from the current state of affairs, as we prepare to *enter* May 2020 with 35, 403 cases, almost thrice the value predicted for the *end* of May. A different model [11] predicted that 54 days of total lockdown would yield around 5, 000 cases. On the 40*^th^* day of lockdown, with seven times as many cases, we can see that this model is not offering good predictions either. Yet another mathematical model [12] estimates that with lockdown from 25*^th^* March onwards (as happened), we would have just shy of 10,000 cases by end-April, which, once again, is one-third the total number of cases right now, and the number of cases by the end of April 25, 2020 was about 2.7 times as predicted.

In the present article, we propose a mathematical model with a different approach, which has been yielding consistent results in predicting the spread since the beginning of the outbreak in India. Moreover, while there exist data-driven approaches to look at the outbreak of COVID-19 [13], [14], there have been no such studies regarding India yet, and this article attempts to fill in that gap as well.

### 4.2. Change Factor

We note that even though the curve of the spread looks exponential, there are daily changes dependent more on the local context — the days immediately preceding. With somewhat lesser weightage but still important is a more global context of the last couple weeks and the pattern in that change. Moreover, once the curve has flattened out and the number of cases have started decreasing, there needs to be a measure for the model to automatically reflect this and adjust predictions accordingly. To account for all of this, we first define a ‘change factor’ — a measure of the ratio of the daily increase with respect to the past *N* days. We start looking at the cases right from January 30, 2020, the day the first COVID-19 case was confirmed in India. The pseudocode for change factor calculation is as in Algorithm 1.

#### Algorithm 1 Calculate change factor for last *N* days with list of daily cases *L*

*sum_change* ← 0

**for** *past day number* = 1 … *N* **do**

*change* ← *L*[*day* – *past day number*]

*sum_change* ← *sum_change + change*

**end for**

*average_change* ← *sum change*/*N*

*actual_change* ← *L*[*day*]

*change_factor* ← *actual_change*/*average_change*

### 4.3. Prediction Based on Raw Data

We have analysed the effects of the pattern in the change factor, the first derivative of cases (i.e., the daily increase, for all of confirmed, deceased and recovered), as well as the second derivative (i.e., the increase in daily increases, for all of confirmed, deceased and recovered) for the raw data in the effectivity of predicting the number of total cases for the following day(s), for confirmed, recovered, as well as deceased cases.

Based on these studies, we came up with Algorithm 2 for predicting the number of daily cases. Here, *α* is a factor we can tune as necessary.

#### Algorithm 2 Predict number of cases for the next day based on last *N* days’ cases, and last *T* days’ change factors with list of daily cases *L*

*trendline* :: change factors for last *T* days

*predicted_factor* ← *sum*(*trendline*)*/T*

*base_cases_considered* ← *sum*(cases in last *N* days)

*base_average* ← *base_cases_considered/N*

*predicted_value* ← *α × predicted_factor* × *base-average*

Extensive experimentation with various values of *α, N* and *T* have yielded that it is most beneficial to currently keep *α* = 1.35, *N* = 3 and *T* = 10. This implies that the predicted cases for the next day is a function of that in the last 3 days, but with an ‘increase’ factor influenced by the last 10 days. This is a testament to and goes at par with the highly dynamic nature of the epidemic.

### 4.4. Prediction Based on Moving Averaged Data

We notice sharp spikes and troughs in the number of daily cases — and this can be due to a variety of reasons, including staggered testing, staggered collection of results, holidays, one-time breakout events, mass gatherings, new testing development, new medical guidelines, and so on. Due to this reason, we also do an analysis of the number of cases (confirmed, recovered and deceased) for a simple moving average of the data over a window of the last *K* days. We have experimented with various values of *K* and have settled on *K* = 3 as the most balanced point wherein we are not diluting the effects of every day’s changes much, but also somewhat accounting for day-to-day external changes in testing and other reasons that are unrelated to the ‘true’ spread of the disease.

With the window averaged list of *K* = 3 days, we use a prediction system the same as in *Algorithm* 2. For the averaged data, the best results were found with an *α* = 0.65 and the same *N* = 3 and *T* = 10, as was found for the prediction with raw (original, unaveraged) data.

### 4.5. Final Prediction

Because the ‘true’ spread of the epidemic is as hard to estimate as the number of cases (for total confirmed, recovered and deceased) per day, we have decided to instead offer a range of predicted values, with one end being the prediction obtained from the raw data and the other end being that obtained from moving averaged data. Over the entire course of time since the outbreak started in India, the model has been 83% accurate in predicting the total number of cases the next day, correctly predicting the range in 74 out of the 90 days looked at. Figure 1 depicts the plot of the actual versus predicted number of daily cases so far, as of May 01, 2020.

**Figure 1:**
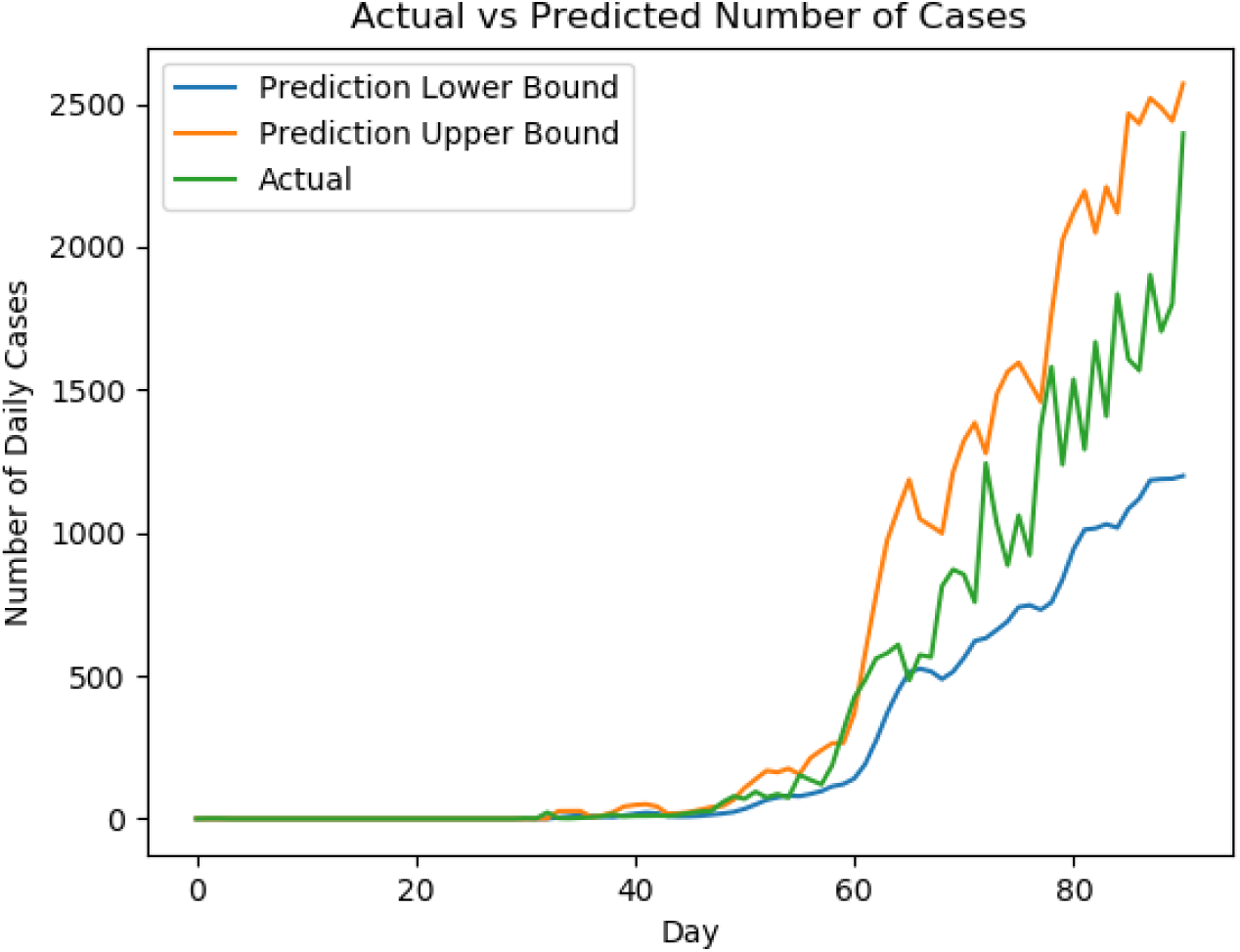
Comparing the Actual and Predicted Number of Daily Confirmed Cases

## 5. Results and Analysis

We observe that the prediction range offered by the raw and moving averaged data offers a high accuracy of 83% until the date of April 30, 2020. We further note, that there is a tradeoff in selecting the values of *α* to be too high or too low — a value much further from 1 will spread out the range and thus have a greater chance of being ‘correct’, but at the same time a huge range prevents a model from being very precise, although accurate. Hence, we have kept the tradeoff balance by predicting a range whose midpoint, on average, deviates only by 61 cases from the actual or true value. This is a mere 3.21% of the maximum daily cases reported in this period, and just 3.39% of the latest day (April 30, 2020)’s reported cases (Figure 2). Moreover, there has been only *one* mis-predicted day in the last 30 days (i.e., over the month of April), with the model thus having 96.67% accuracy over the last month. We note, most of the prediction errors are around days 30 to 42. The model is self-correcting as it improves its predictions every day, by incorporating the previous day’s data into the trendline for the following days. This model can thus be used dynamically not only to predict the spread of COVID-19 in India, but also to check the effect of various government measures in a short span of time after they are implemented.

**Figure 2:**
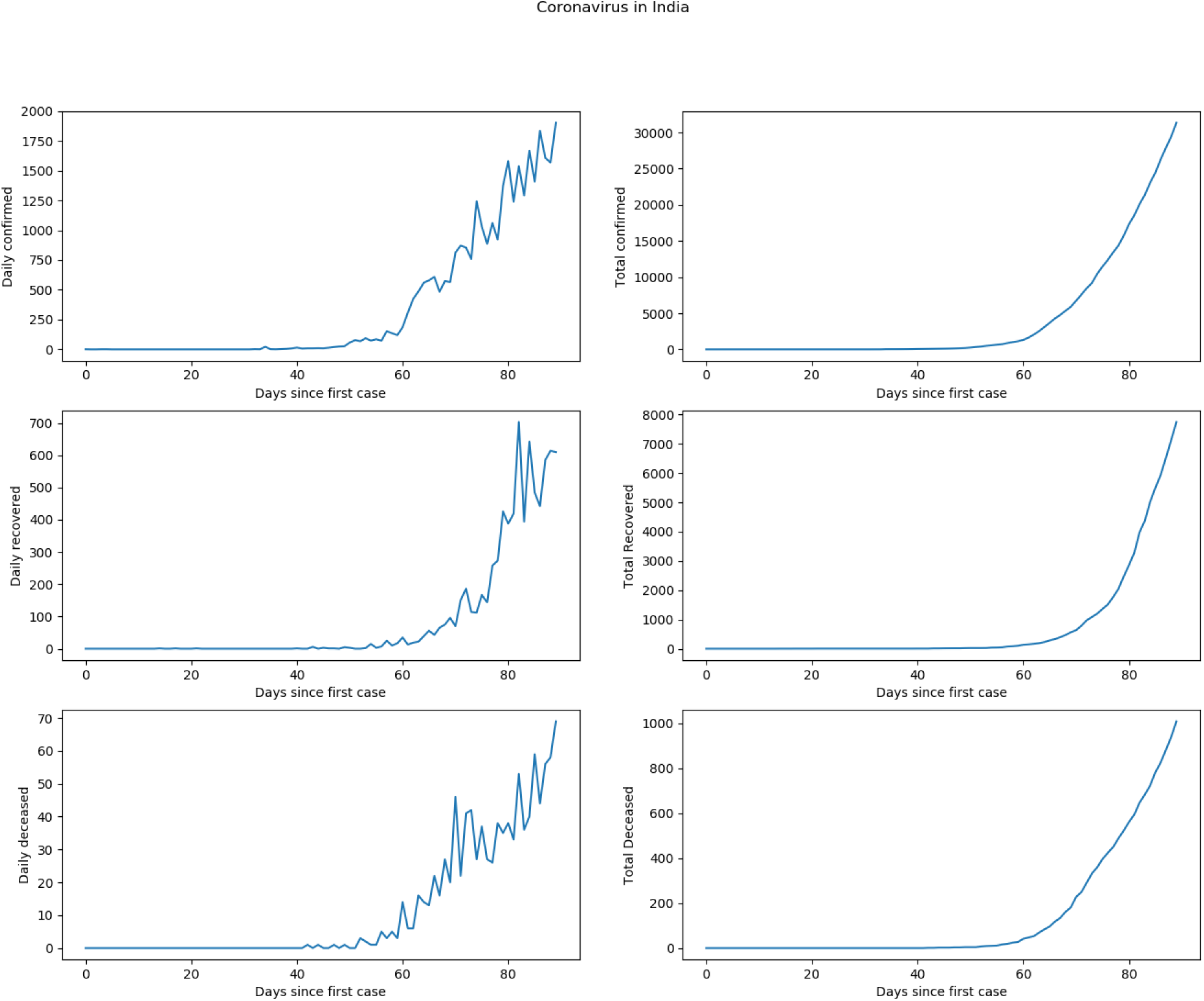
COVID-19 Cases in India

## 6. Conclusion and Future Work

We can see an alarming growth in the number of cases in India over the last month, but also a silver lining in the current ‘stabilising’ nature of trend factors, possibly owing to the lockdown measures implemented, along with widespread testing, contact tracing, and general hygiene measures. Moreover, we also notice that a lot of the sharp spikes and new infections are for mass gathering events and the close contacts of the infected, respectively, highlighting even more the need for physical distancing and maintaining good hygiene. The age distribution among the known confirmed cases in India show a tilt towards the younger population, a departure from the world trend. The quick turnaround of recovery in infected patients in India is another beacon of hope.

Further work is being done to incorporate more factors into the model and break it down statewise to gain more insight into the status of the spread in various states of India and fine-tune it to the needs of each individual state. We also hope to modify the present model to adapt it to the needs of different countries in the future. Additional work includes the analysis of the change factors. Brief investigation into the list of change factors shows that a factor that stays around 1 indicates a ‘stabilization’ in the daily number of confirmed cases.

## Data Availability

All the data referred to in the manuscript are open-sourced.

https://api.covid19india.org/

https://www.mohfw.gov.in/

## Supplement: Important Figures and Plots

Attached as supplement are some important figures that helped us identify trends and deduce conclusions. They offer a great insight into the current spread in the country.

Figure 2 shows us the number of daily as well as total confirmed, recovered and deceased COVID-19 cases in India. Notably, there are a lot of peaks and troughs in the daily cases’ graphs. To account for possible external factors unrelated to true spread of the disease that are causing these spikes, we have Figure 3, a simple moving averaged data with the window size *K* = 3. We note the smoother nature of the curves depicting daily cases in this figure.

**Figure 3:**
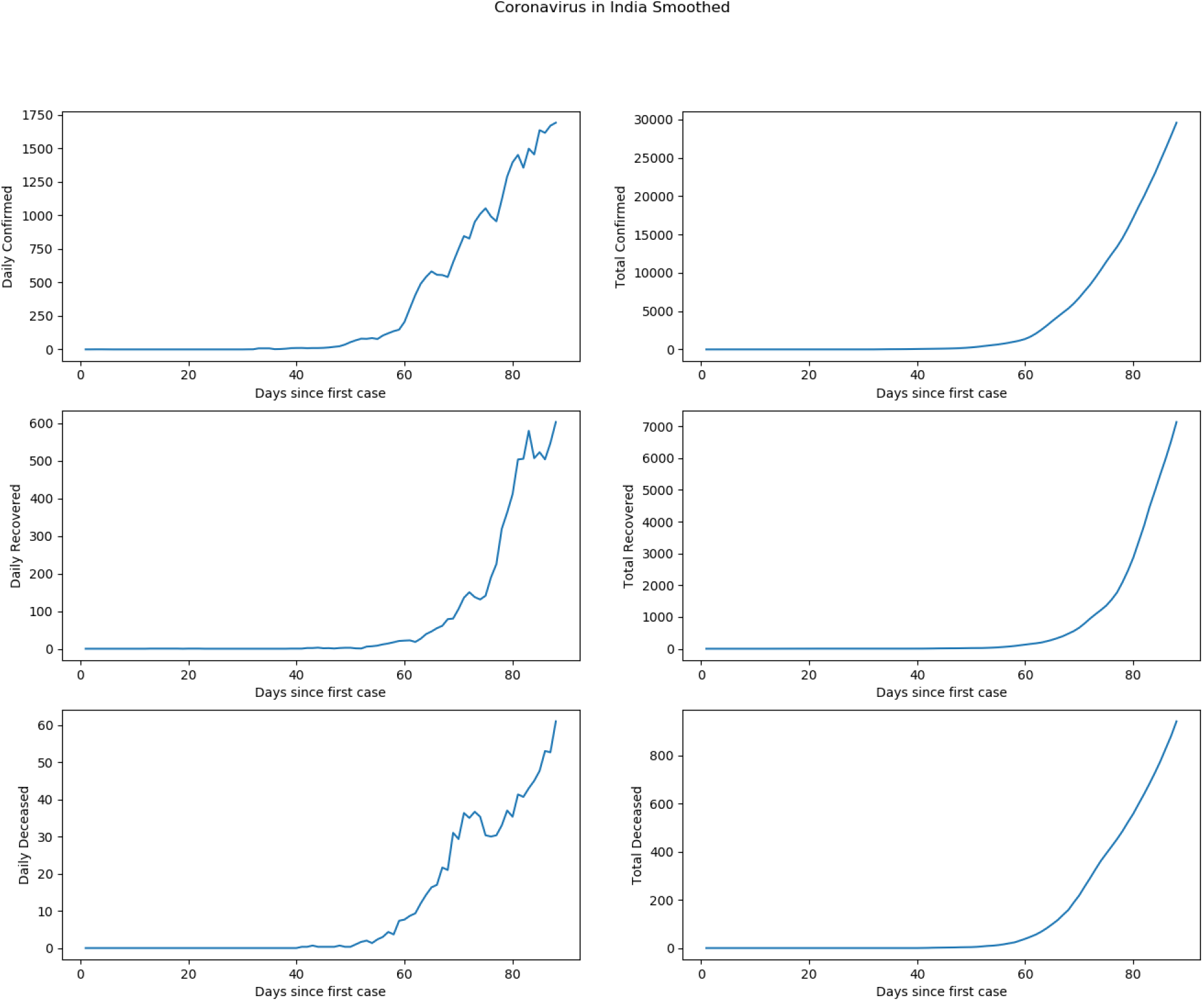
COVID-19 Cases in India, Moving Averaged with *K* = 3

Figure 4 denotes the same data as Figure 3 does, but on a logarithmic scale. Realizing the exponential nature of this pandemic, we have decided to include this log-scale plot. A straight line on a logarithmic scale indicates an exponential rise in the absolute number of cases. Looking at the three subplots in Column 2 of Figure 4, we note this alarming trend.

**Figure 4:**
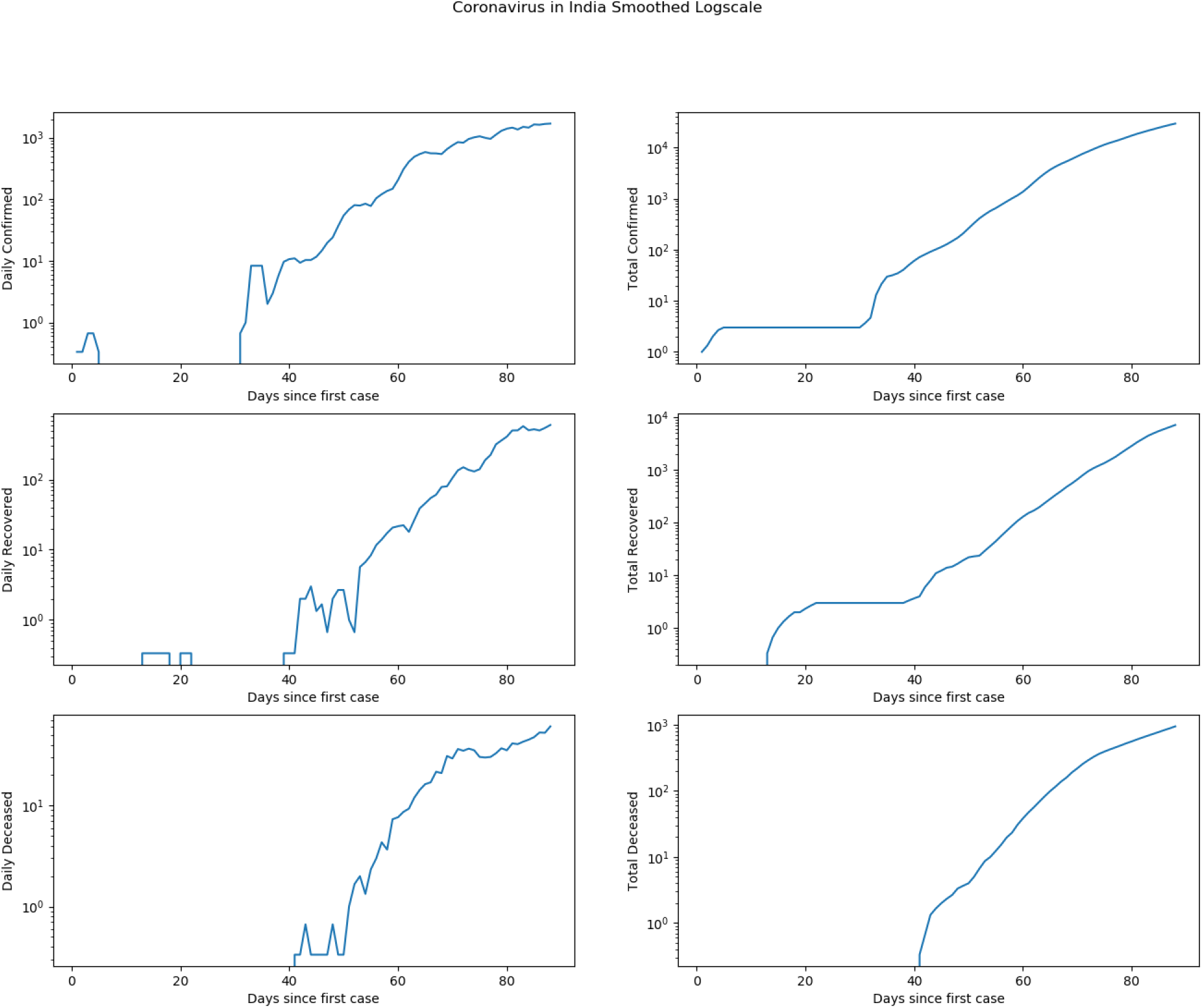
COVID-19 Cases in India on a Log Scale, Moving Averaged with *K* = 3

Finally, in Figure 5, we plot the corresponding derivatives of the subplots in Figure 3. The three subplots in Column 1 denote the first derivative of number of *daily* cases, i.e., how the number of daily new cases is changing. We see sharp large fluctuations towards the end of the subplots, indicating the difficulty that any predictive system faces. In Column 2, we have the first derivative of the *total* number of cases, i.e., the number of daily cases. We see the similarity between these plots to those in Column 1 of Figure 3, as the derivative of the cumulative total is nothing but the number of daily cases.

**Figure 5:**
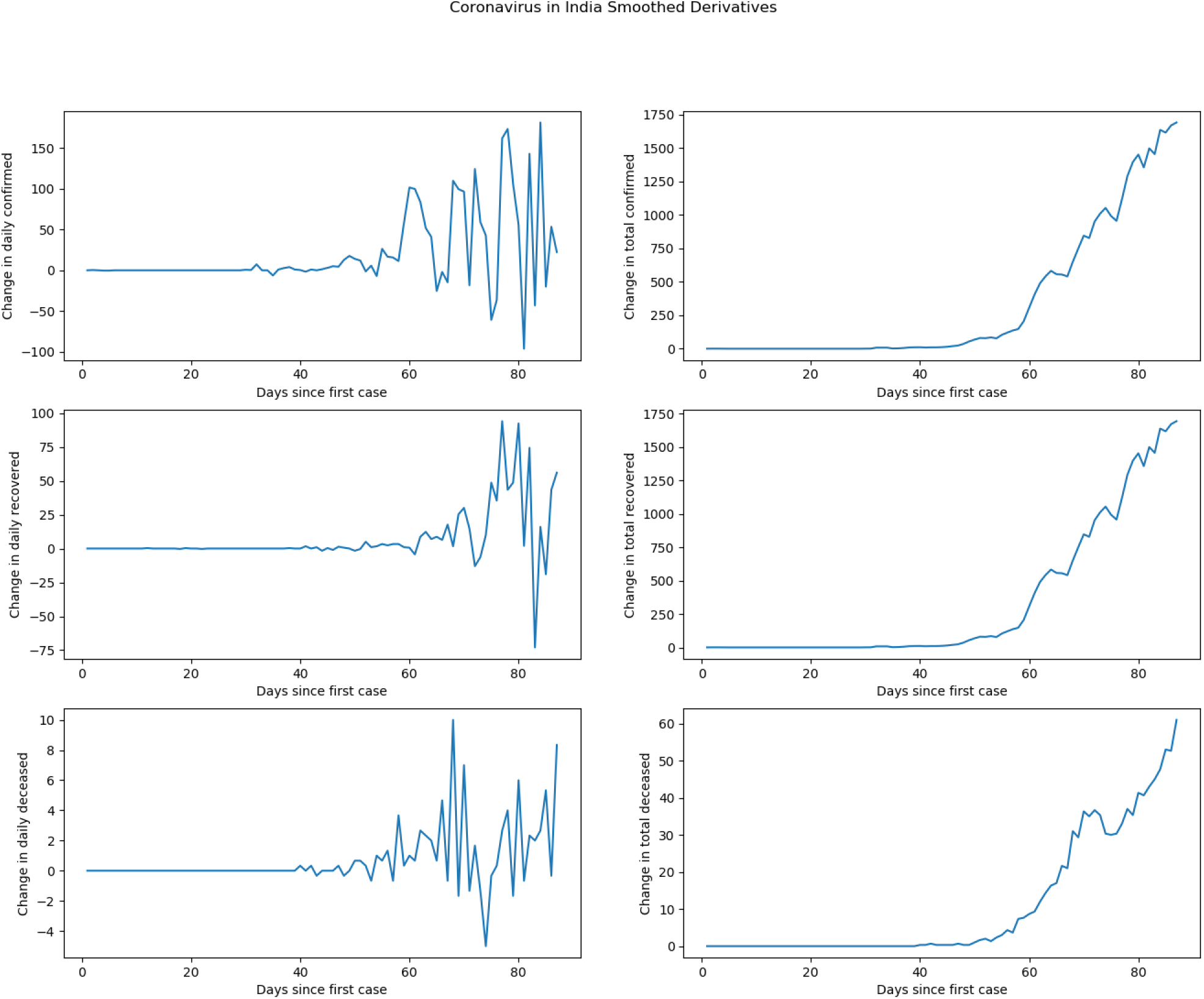
First Derivative of COVID-19 Cases in India, Moving Averaged with *K* = 3

## References

[1] https://www.who.int/emergencies/diseases/novel-coronavirus-2019 (accessed April 28, 2020).

[2] https://www.mohfw.gov.in/ (accessed April 28, 2020).

[3] https://www.who.int/docs/default-source/wrindia/india-situation-report-1.pdf?sfvrsn=5ca2a672_0 (accessed April 28, 2020).

[4] https://www.worldometers.info/coronavirus/ (accessed April 28, 2020).

[5] https://www.statista.com/statistics/1103023/coronavirus-cases-distribution-by-age-group-italy/ (accessed April 28, 2020).

[6] http://predictivehealthcare.pennmedicine.org/2020/03/14/accouncing-chime.html (accessed May 1, 2020).

[7] https://hcdatascienceservices.com/COVID19CapPlan/?visit=Content-Mask-1.7.0.11#application_guidance (accessed May 1, 2020).

[8] I. Ciufolini, A. Paolozzi, Mathematical prediction of the time evolution of the covid-19 pandemic in italy by a gauss error function and monte carlo simulations, Eur. Phys. J. PlusarXiv:https://link.springer.com/article/10.1140/epjp/s13360-020-00383-y, doi:10.1140/epjp/s13360-020-00383-y.

[9] https://www.cdc.gov/coronavirus/2019-ncov/covid-data/forecasting-us.html (accessed May 1, 2020).

[10] R. Ranjan, Predictions for covid-19 outbreak in india using epidemiological models, medRxivarXiv:https://www.medrxiv.org/content/early/2020/04/12/2020.04.02.20051466.full.pdf, doi:10.1101/2020.04.02.20051466.

[11] A. M.K., Modeling and predictions for covid 19 spread in indiadoi:10.13140/RG.2.2.11427.81444.

[12] S. Roy, K. Roy Bhattacharya, Spread of covid-19 in india: A mathematical model doi:10.13140/RG.2.2.15878.52802.

[13] Y. Fang, Y. Nie, M. Penny, Transmission dynamics of the covid-19 outbreak and effectiveness of government interventions: A data-driven analysis, Journal of Medical Virology 92 (6) (2020) 645–659. arXiv:https://onlinelibrary.wiley.com/doi/pdf/10.1002/jmv.25750, doi:10.1002/jmv.25750. URL https://onlinelibrary.wiley.com/doi/abs/10.1002/jmv.25750

[14] S. K. Dey, M. M. Rahman, U. R. Siddiqi, A. Howlader, Analyzing the epidemiological outbreak of covid-19: A visual exploratory data analysis approach, Journal of Medical Virology 92 (6) (2020) 632–638. arXiv:https://onlinelibrary.wiley.com/doi/pdf/10.1002/jmv.25743, doi:10.1002/jmv.25743. URL https://onlinelibrary.wiley.com/doi/abs/10.1002/jmv.25743

